# Demographic and clinical differences in patients with positional obstructive sleep apnea and development of a discrimination model

**DOI:** 10.1101/2020.07.28.20164053

**Authors:** Shang-Yang Lin, Cheng-Yu Tsai, Wen-Te Liu

**Author notes:** Corresponding Author: Wen-Te Liu. E-mail: Shang-Yang Lin, Cheng-Yu Tsai.

## Abstract

**Purpose:** Obstructive sleep apnea (OSA) is a highly prevalent disease, and positional OSA (pOSA) is a subgroup whose OSA severity is highly affected by sleeping position. This study investigated differences in demographic and sleep characteristics between patients with and without pOSA and developed a simple discrimination model.

**Methods:** We reviewed polysomnography records of patients admitted to the Sleep Center at Shuang-Ho Hospital between March 2015 and March 2019. They were categorized into pOSA and non-pOSA groups, and their demographic and sleep characteristics were compared. The receiver operating characteristic (ROC) curve was used to estimate the feasibility of discrimination model.

**Results:** Of the patients, 33% received diagnoses of pOSA; they had smaller neck circumference and waistline and lower weight, body mass index (BMI), OSA severity, heart rate, and snoring and respiratory-related limb movement indexes but higher sleep efficiency and mean oxygen saturation compared with patients without pOSA. Sleep stage analysis revealed that as severity increased, the proportion of sleep time spent in N2, N3, and rapid eye movement stages decreased, but the proportion of time spent in the N1 stage increased in both populations. Sleep position analysis revealed a higher proportion of sleep time in a supine position among patients with pOSA after adjustment for severity. The corresponding area under the ROC curve of our discrimination model was 0.924.

**Conclusions:** Demographic and sleep characteristics differed significantly between patients with and without pOSA. Our model uses readily available measurements such as BMI and waistline and can aid physicians in the timely identification of patients with pOSA.

**Trial registration number:** TMU-JIRB No.: N201911007

**Date of registration:** 2019/11/12

## Introduction

Obstructive sleep apnea (OSA) is a common sleep disorder characterized by repeated episodes of apnea during sleep, often in the form of brief awakenings due to complete or partial upper airway obstruction causing blood oxygen desaturation [1]. Obesity and aging are two major risk factors for OSA, which is becoming a crucial public health concern in modern society [2, 3]. Growing evidence has revealed that sleep disorders not only influence academic and work performance, increase the risk of dementia, and worsen depression but also adversely affect multiple organs and systems and aggravate cardiovascular diseases, respiratory diseases, metabolic dysfunction, cognitive decline, and even the progression of cancer [4–7]. Because Asian populations tend to have a narrower cranial base and flatter midface structure compared with those of Caucasian populations, they are estimated to have a greater prevalence of sleep disorders [8].

Positional OSA (pOSA) is a subgroup of OSA that is defined as an apnea hypopnea index (AHI) in a supine position that is at least twice as high as that in a nonsupine position but for which the AHI in a nonsupine position is less than 5 [9]. Studies have suggested that position changes during sleep may affect sleep quality, sleep architecture, and even sleepiness levels during daytime [10, 11]. Furthermore, the supine position was proven to be prone to sleep apnea events [12, 13].

Traditional positional therapy (PT) for preventing patients from sleeping in a supine position involved the attachment of a tennis ball between the patient’s shoulder blades, and this was effective for reducing the amount of time spent in a supine position during sleep, ameliorating OSA symptoms, and preventing sleepiness during daytime and oxygen desaturation [17]. One study conducted in Korea reported a prevalence of pOSA of up to 75.6% [18]. However, comprehensive investigations focusing on the demographic and sleep characteristics of individuals with pOSA are scarce, especially in Asian countries.

Customized therapy strategies are difficult for physicians to design without an accurate diagnosis. However, in recent years, the increasing awareness of sleep disorders has not corresponded with an increase in the number of beds available for sleep monitoring; consequently, waiting times for hospital-based polysomnography (PSG) monitoring are continually increasing [19]. Portable devices designed for home sleep tests may be a time- and cost-efficient alternative; moreover, these devices are considered to be more credible because they more closely reflect real situations [20]. Of course, a discrimination model for identifying patients with pOSA by using only measurements that can be easily obtained with simple tools or portable devices would be useful for diagnosis and treatment strategy formulation; such a model could also mitigate the individual, hospital, and government burden of sleep medicine.

OSA is a critical disorder, and it can cause other complications or evolve into a critical disease without appropriate treatment. To improve knowledge on pOSA and achieve the timely identification of affected patients, which would assist physicians in formulating improved program treatment strategies in clinical settings, we retrospectively reviewed the PSG records of patients who were referred to the Sleep Center of Shuang-Ho Hospital, New Taipei City, Taiwan (ROC). This study had two objectives: the first was to investigate the prevalence of pOSA as well as the demographic features and clinical characteristics of affected patients referred to Shuang-Ho Hospital. The second was to develop a method for the accurate and simple identification of patients with pOSA. Accordingly, we identified significant factors and then used these factors to construct a discrimination model.

## Materials and Methods

The Institutional Review Board of Taipei Medical University approved the retrospective analysis of a clinical sleep center database without requiring additional consent (TMU-JIRB No.: N201911007). We reviewed the medical records of 7131 individuals who underwent clinical PSG at the Sleep Center of Shuang-Ho Hospital between March 2015 and March 2019. Subsequently, after patients without complete records and those without OSA (AHI ≤ 5) were excluded, 4440 patients with OSA were included in the final analyses. Patients with OSA were designated to either the pOSA or non-pOSA group according to the following definition of pOSA proposed by Mador: patients whose AHI in a supine position was at least twice as high as that in a nonsupine position and whose nonsupine AHI was less than 5 [9].

Two groups were compared in terms of demographic features, including height, weight, body mass index (BMI), age, neck circumference, and waistline, as well as the clinical characteristics obtained through PSG, including total sleep time, sleep efficiency, wake after sleep onset (WASO) period, AHI, respiratory disorder index (RDI), mean and minimum oxygen saturation, desaturation index, heart rate, and snoring and respiratory-related limb movement indexes. The distribution normality of each parameter was evaluated using the Shapiro–Wilks test, and then Student’s *t* test or the Mann–Whitney *U* test was employed according to the data type. Fisher’s exact test was conducted to examine the difference in gender composition between the two populations. For the analysis of sleep architecture and position, to minimize the influence of severity, individuals in each group were further categorized into three groups according to OSA severity: mild (5 ≤ AHI < 15), moderate (15 ≤ AHI < 30), and severe (AHI ≥ 30). Because sleep stage and posture were presented as proportions of total sleep time, Kruskal–Wallis and Bonferroni post hoc tests were used for sleep architecture and posture analysis.

When developing the discrimination model, we randomly selected and integrated 50% of the data in each group to form one data set for determining the importance of each factor. Multivariate logistic regression with stepwise selection was employed to develop a model for identifying patients who met the criteria for pOSA. Within this regression equation, 27 factors, including demographic features and clinical characteristics, were input as independent variables. The remaining data were used to validate our model. Sensitivity, specificity, and accuracy were calculated for our model, and discrimination ability was estimated according to the area under the receiver operating characteristic (ROC) curve (AUC). The probability threshold used for OSA classification was 0.50. All results were obtained using R software.

## Results

### Prevalence and demographic features of pOSA

Nearly one-third of the patients (1102 patients, 33.01%) were diagnosed as having pOSA; they had a lower weight (*p* ≤ 0.001), lower BMI (*p* ≤ 0.001), and smaller neck circumference and waistline (both *p* ≤ 0.001) than did those without pOSA (Table 1). The proportion of female patients was 30.94% in the pOSA population but only 19.59% in the non-pOSA population (Fisher’s exact test: *p* < 0.001).

**Table 1.** Demographic features and clinical characteristics of patients with and without positional obstructive sleep apnea (pOSA). Abbreviations: BMI: body mass index, WASO: wake after sleep onset, AHI: apnea hypopnea index, RDI: respiratory disturbance index, RRLM: respiratory-related limb movements. Units for parameters are presented in parentheses (if applicable).

|  | pOSA | Non-pOSA | Statistics |
| --- | --- | --- | --- |
| **Demographic features** |  |  |  |
| Height (cm) | 166.10 ± 9.02 | 167.81 ± 8.23 | $p < 0.001$ |
| Weight (kg) | 70.77 ± 13.00 | 81.24 ± 16.07 | $p < 0.001$ |
| BMI | 25.63 ± 4.98 | 28.77 ± 4.91 | $p < 0.001$ |
| Age (yr) | 50.37 ± 13.20 | 49.54 ± 12.88 | $p = 0.53$ |
| Neck circumference (cm) | 37.40 ± 10.50 | 40.20 ± 10.60 | $p < 0.001$ |
| Waistline (cm) | 88.44 ± 27.66 | 97.18 ± 12.65 | $p < 0.001$ |
| **Clinical characteristics** |  |  |  |
| Total sleep time (hr) | 5.69 ± 0.49 | 5.70 ± 0.50 | $p = 0.58$ |
| Sleep efficiency (%) | 78.11 ± 15.04 % | 75.64 ± 15.90 % | $p < 0.001$ |
| WASO (min) | 56.45 ± 46.61 | 66.65 ± 48.99 | $p < 0.001$ |
| AHI | 18.59 ± 15.03 | 42.27 ± 25.76 | $p < 0.001$ |
| RDI | 18.85 ± 15.04 | 42.46 ± 25.71 | $p < 0.001$ |
| Mean oxygen saturation (%) | 96.19 ± 1.58 % | 94.77 ± 2.44 % | $p < 0.001$ |
| Oxygen desaturation index | 17.96 ± 15.21 % | 41.29 ± 26.07 % | $p < 0.001$ |
| Desaturation arousal index | 1.46 ± 2.20 | 3.66 ± 4.25 | $p < 0.001$ |
| Heart rate (bpm) | 65.52 ± 8.65 | 68.05 ± 9.79 | $p < 0.001$ |
| Snoring index | 209.41 ± 199.20 | 284.31 ± 216.72 | $p < 0.001$ |
| RRLM index | 1.86 ± 3.40 | 4.77 ± 6.48 | $p < 0.001$ |

### Clinical characteristics of pOSA

Regarding sleep characteristics, PSG revealed that patients with pOSA had a lower AHI (*p* < 0.001), slower heart rate, lower snoring and respiratory-related limb movement indexes (all *p* < 0.001), and higher sleep efficiency and mean oxygen saturation (both *p* ≤ 0.001) compared with those in the non-pOSA group (Table 1). On the basis of OSA severity, we further separated each population into three subgroups: mild (5 ≤ AHI < 15), moderate (15 ≤ AHI < 30), and severe (AHI ≥ 30). The two populations exhibited distinct compositions: the pOSA population was mainly composed of patients with mild OSA, and the non-pOSA population had a high proportion of patients with severe OSA. The proportions of patients with mild, moderate, and severe OSA were respectively 56.53%, 28.31%, and 15.15% in the pOSA population and 14.50%, 23.76%, and 61.74% in the non-pOSA population (Fig. 1). Sleep stage analysis revealed a clear trend in both populations that as severity increased, the amount of time spent in the N2, N3, and rapid eye movement stages decreased, and that in the N1 stage increased (Table 2). Sleep position analysis revealed a higher supine ratio in all three severity subgroups in the pOSA population than in the non-pOSA population (all *p* < 0.001) (Table 2).

**Fig. 1:**
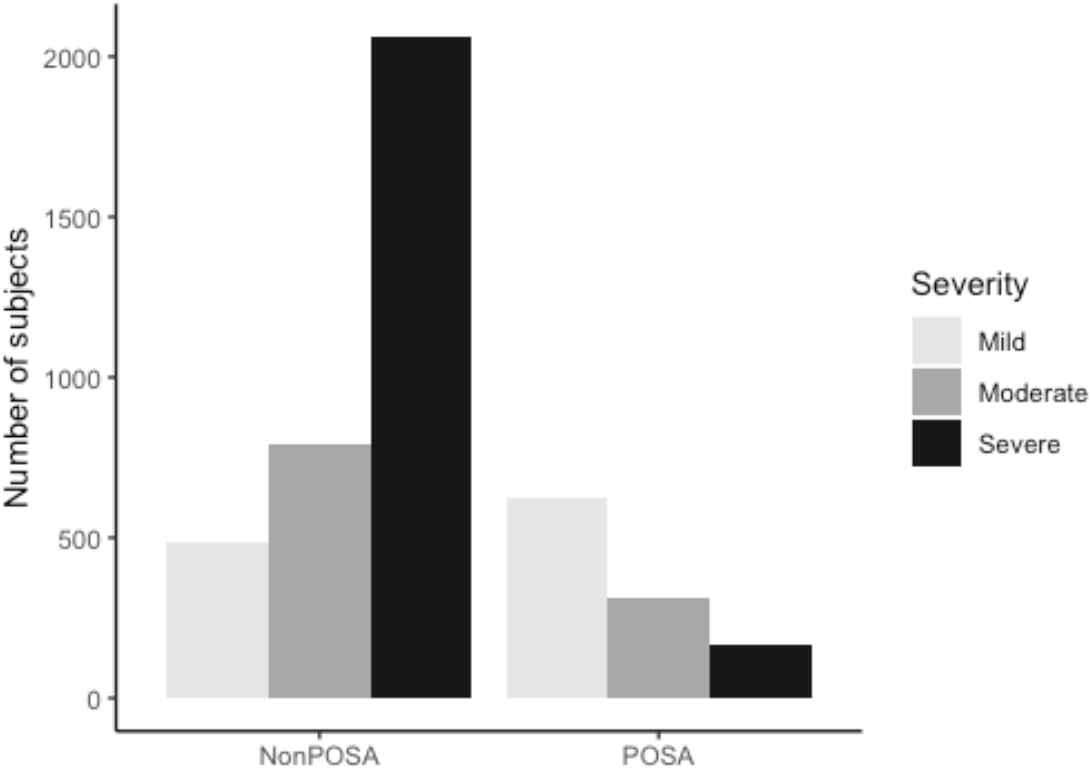
Number of patients in each category of OSA severity in the pOSA and non-pOSA groups. The compositions of the pOSA and non-pOSA groups were significantly different.

**Table 2.** Proportion of patients with each sleep stage and sleep position in subgroups categorized according to OSA severity and type.

|  | pOSA |  |  | Non-pOSA |  |  | Statistics |
| --- | --- | --- | --- | --- | --- | --- | --- |
|  | Mild | Moderate | Severe | Mild | Moderate | Severe |  |
| N | 623 | 312 | 167 | 484 | 793 | 2061 |  |
| Proportion | 56.53% | 28.31% | 15.15% | 14.50% | 23.76% | 61.74% |  |
| **Sleep stages** |  |  |  |  |  |  |  |
| N1 | 11.29% | 13.12% | 17.90% | 12.00% | 14.54% | 20.28% | $p < 0.001$ |
| N2 | 70.20% | 70.13% | 68.04% | 70.44% | 68.28% | 66.14% | $p < 0.001$ |
| N3 | 4.95% | 3.67% | 2.57% | 4.24% | 3.86% | 2.29% | $p < 0.001$ |
| REM | 13.56% | 13.09% | 11.50% | 13.32% | 13.31% | 11.29% | $p < 0.001$ |
| **Sleeping position** |  |  |  |  |  |  |  |
| Supine proportion | 64.53% | 74.35% | 88.09% | 48.62% | 52.15% | 58.57% | $p < 0.001$ |

### Prediction model construction and validation

Multivariate logistic regression with stepwise selection indicated that six factors were significant in determining whether an individual met the criteria of pOSA: BMI, waistline, mean oxygen saturation, total sleep time, severity of OSA, and proportion of time spent in a supine position during sleep (Table 3). The coefficients revealed that mean oxygen saturation percentage and proportion of time spent in a supine position during sleep were positive predictors, whereas BMI, waistline, total sleep time, and OSA severity were negative predictors. The six significant factors were used to construct the discrimination model, which achieved a sensitivity of 100% and specificity of 75.3%. The corresponding AUC was 0.924, indicating excellent discrimination ability of the model with six easily obtained parameters (Fig. 2).

**Table 3.** Multivariate logistic regression analysis revealed six significant factors for identifying individuals with pOSA. Abbreviation: BMI: body mass index.

| Variable | $\beta$ | Std. error | *p* |
| --- | --- | --- | --- |
| BMI | −0.02 | 0.003 | <0.001 |
| Waistline | −0.001 | 0.001 | 0.03 |
| Mean oxygen saturation (SaO2) | 1.59 | 0.448 | <0.001 |
| Total sleep time | −0.03 | 0.013 | 0.02 |
| Severity of OSA | −0.50 | 0.017 | <0.001 |
| Supine ratio | 0.21 | 0.027 | <0.001 |

**Fig. 2:**
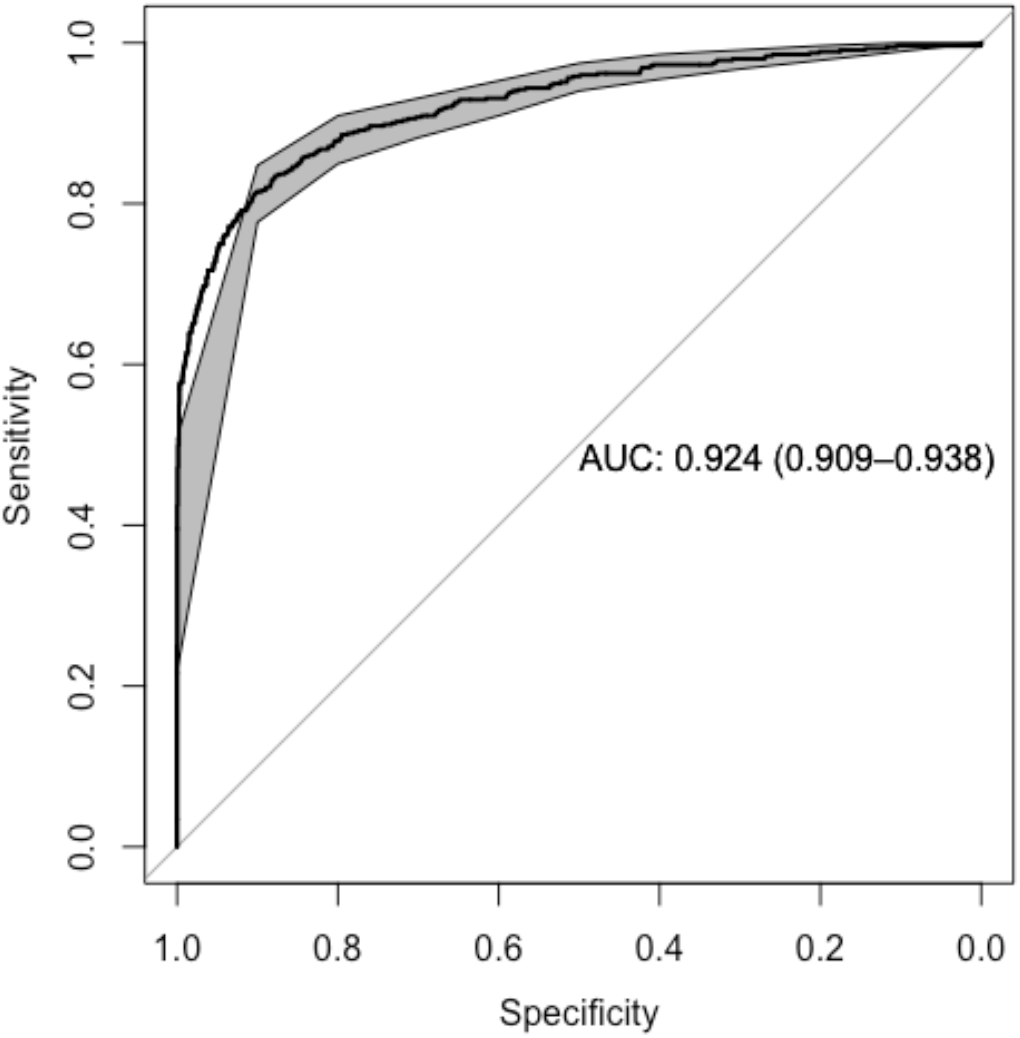
AUC of 0.924, indicating excellent ability to differentiate between patients with and without pOSA.

## Discussion

Patients with pOSA comprise a subgroup of those with OSA whose OSA severity is highly affected by their sleeping position. Specifically, patients with pOSA were prone to experiencing apnea while sleeping in a supine position but not while sleeping in other positions. Although some agreement has been reached regarding the basic concept of pOSA, controversy remains regarding its prevalence. Cartwright defined pOSA as a supine AHI at least twice the nonsupine AHI [21]. Mador used the same criterion with the addition of nonsupine AHI < 5 [9]. The Dutch criteria are the same as Mador’s, with the addition that the proportion of time spent in a supine position during sleep must be between 10% and 90% of the total sleep time to avoid influence from inaccurate extreme values [22]. Previous investigations have reported considerably varied pOSA prevalence ranging from 27.7% to 75.6% [18, 23, 24]. A study conducted in Denmark that involved the comparison of various inclusion criteria in a single data set concluded that the prevalence of pOSA depends fundamentally on the definition used [25].

Despite indicating a different prevalence, our data agreed with those of investigations conducted in Chile, Turkey, and Korea with respect to most demographical features and clinical characteristics of patients with pOSA. Compared with the non-pOSA group, the pOSA group had lower AHI values, a high proportion of women, lower BMI values, and smaller neck circumferences [18, 23, 24]. Studies have also proposed that lower sleepiness levels and prevalence of hypertension or other diseases among patients with pOSA can be attributed to the lower AHI of these patients compared with those without pOSA [23, 24]. Similarly, patients with pOSA had slower heart rates, lower values for snoring and respiratory-related limb movement indexes, greater sleep efficiency, and higher mean oxygen saturation levels. Lee and colleagues proposed that patients with pOSA tend to be older than those without [18], but no statistically significant difference was observed in our data. Moreover, the results obtained from sleep position analysis were different; our data indicated that the amount of time spent sleeping in a supine position increased with OSA severity (in both pOSA and non-pOSA groups) and was greater in the pOSA group than in the non-pOSA group in all severity subgroups, but the aforementioned study conducted in Korea reported an opposing trend [18].

Obesity was a main component contributing to sleep apnea [3]. Sleep duration and quality decreased with increased body weight and adiposity [26, 27]. Although the evidence was less direct, OSA also reciprocally contributed to obesity [28]. In addition, studies have demonstrated that OSA adversely affects multiple organs and systems, with particular relevance to cardiovascular diseases [7, 8]. These may be triggered by intermittent hypoxemia, which was a characteristic of patients with OSA; moreover, it promotes oxidative stress, increases sympathetic activation and blood pressure, drives systemic and vascular inflammation, and causes diverse multiple organ chronic morbidity and mortality associated with cardiovascular disease, metabolic dysfunction, cognitive decline, and even the progression of cancer [29]. According to the demographic and clinical characteristics of patients with pOSA, we speculate that patients with pOSA were in the early stages of OSA. Effectively reducing the amount of time spent sleeping in a supine position and executing weight management can ameliorate symptoms and reduce the risk of developing critical illnesses, including cognitive impairment and chronic renal disease.

Currently, the therapy most recommended to patients with OSA is continuous positive airway pressure, but lack of compliance due to discomfort detracts from its beneficial effects. PT, which aims to prevent patients from sleeping in a supine position, is cost effective and is categorized into three main types according to the materials used: a simple tennis ball, a battery-powered positioning device, and a special-shaped pillow [30]. With effective reductions in the proportion of time spent sleeping in a supine position, PT was shown to ameliorate symptoms of OSA and prevent sleepiness during the daytime and oxygen desaturation without affecting sleep architecture [17]. PT also boasts high compliance rates and long-term efficacy [17]. Moreover, studies have indicated that patients without obesity are more likely to benefit from changes in sleeping position [13, 21], which underscores the importance of early therapy for patients with pOSA.

Prolonged waiting times for hospital-based PSG can delay treatment and deter patients from complying with sleep studies and subsequent treatment. Therefore, the development of alternative diagnostic tools that are simple and highly accurate in identifying patients with pOSA is urgently required and will facilitate the development of patient-specific treatment strategies. This study demonstrated the possibility of developing a pOSA discrimination model by using readily available measurements that can be obtained using simple tools or portable sleep-monitoring devices. Our model demonstrated outstanding ability in discriminating patients with pOSA from those with non-pOSA as well as high sensitivity and specificity. Our next steps will be validating the developed model by using a multihospital database and comparing computing model accuracy between the use of data obtained through PSG and the use of that obtained from portable devices. Our discrimination model provides a rapid, accurate, and reproducible method for predicting whether patients have pOSA. The model may be clinically useful as a screening tool for pOSA rather than as a replacement for PSG.

## Conclusion

Positional OSA groups have distinct demographical features and clinical characteristics, and we speculate that patients with pOSA are in the early stages of OSA. We used readily available measurements to identify patients with pOSA with high accuracy and simplicity, and our findings can aid clinical physicians in the timely formulation of customized treatment programs.

## Data Availability

The data that support the findings of this study are available on request from the corresponding author, Wen-Te Liu. The data are not publicly available due to involving the privacy of research participants.

## Acknowledgments

This study was supported by the Taiwan Ministry of Science and Technology (MOST 108-2634-F-038-003). We thank Hsin-Chien Lee, MD; Ying-Shuo Hsu, MD; and Cheng-Rong Wu, MD for their comments on sleep medicine and all technicians at the Sleep Center, Shuang-Ho Hospital, for their contribution to data collection and processing.

